# EuroQol-5D-3L in Long Covid patients After Supplementation with EchA Marine^®^, a Sea Urchin Eggs Extract: a double-blinded, multicentrical study

**DOI:** 10.1101/2023.05.31.23290798

**Authors:** V Brichetti, T. Rubilar, J. Tejada, P. Montecino, A. Crespi-Abril, E Barbieri, N de la Rosa, J. Iriarte-Vazquez, N. Jajati, C. Volonteri, M. Sivori, G. de Larrañaga, F. Saldarini

## Abstract

Patients with Long COVID experience a significant decrease in their quality of life and the lack of effective treatment represents an unmet need in medical care and patient health. One proposed strategy for treating Long COVID is to increase the body’s ability to restore immune balance by controlling inflammation with anti-inflammatory substances. For this reason, the aim of this double-blind study was to evaluate the supplementation of patients with EchA Marine^®^, a dietary supplement based on sea urchin eggs rich in Echinochrome A. This compound has demonstrated anti-inflammatory and antioxidant properties by activating the metabolism of glutathione and improving mitochondrial mass and performance. The EuroQol 5-Dimension (EQ-5D) is a standardized questionnaire assessing five dimensions of health: mobility, self-care, usual activities, pain/discomfort, and anxiety/depression used as an instrument to measure health-related quality of life in clinical and economic studies. In this multicenter, double-blind, intervention study, we have demonstrated that the dietary supplement EchA Marine^®^ can significantly enhance the quality of life of these patients, particularly in pain and discomfort; notably improving their quality of life and daily activity’s ability. EchA Marine^®^ is an effective treatment option for Long COVID patients and with further research its efficacy could be further strengthened.

## Introduction

The COVID-19 pandemic impacted the health of millions of people worldwide, and the SARS-COV-2 infection can also lead to persistent symptoms lasting for many weeks or even longer. Current literature has not settled the issue of what to call this syndrome and it is known as post-acute sequelae of SARS-CoV-2 infection’ (PASC) and ‘post-acute coronavirus disease (COVID) syndrome’ (PACS), as well as being more widely known as ‘Long COVID’. Due to the fact that the affected individuals continue to experience long-lasting and debilitating symptoms, the syndrome is commonly referred to as Long COVID or “COVID sequelae” ^[1]^. The definition of this Long COVID syndrome is characterized by a number of symptoms that persist over time in individuals who have had COVID-19 beyond 12 weeks from having contracted the disease or from having received their diagnosis, and which symptoms have not been able to be explained by an alternative diagnosis^[2,3,4,5,6]^. These symptoms can include fatigue, difficulty in breathing, headache, loss of smell and taste, and muscle pains, amongst others. Patients with Long COVID experience a significant decrease in their quality of life, making them particularly vulnerable to the various negative effects of Long COVID^[7,8,9]^. To date, the Long COVID syndrome is not fully understood and the lack of effective treatment represents an unmet need in medical care and patient health^[10]^. Efforts have been made to understand the effect of Long COVID on patients’ quality of life as a health-related quality of life assessment is essential to the understanding of COVID-19’s total impact, as well as serving as a guide for clinical and policy decisions.

One commonly used tool to measure health-related quality of life is the EuroQol 5-Dimension (EQ-5D), which is a standardized questionnaire assessing five dimensions of health: mobility, self-care, usual activities, pain/discomfort, and anxiety/depression. The EuroQol is a widely used instrument to measure health-related quality of life in clinical and economic studies. It comprises of a brief and simple questionnaire allowing for the comparison of results between different diseases and treatments^[11]^. Additionally, it includes a visual analog scale (VAS) that measures overall health perception. Based on these attributes, several studies have implemented the EuroQol questionnaire to evaluate the health-related quality of life in patients with Long-COVID^[4,12,13,14,15]^. Overall, the EuroQol has proven to be a valuable tool for assessing quality of life in a wide range of medical conditions and appears to be particularly useful for assessing the quality of life in patients with Long-COVID-19^[9,16,17]^. With specific reference to Long-COVID, the EuroQol has been able to detect the decrease in health-related quality of life and has been able to demonstrate that Long COVID involves a significantly higher disability rate than that which is seen in the general population^[9,18,19,20]^.

In general, COVID-19 pathologies are still marked by a cytokine storm that leads to inflammation of the endothelium, microvascular thrombosis, and multiple organ failures^[21]^. The mechanisms that may be involved in Long COVID may not only be viral reservoirs, but also multi-organ tissue damage, reactivation of neurotrophic pathogens, such us Epstein-Barr, production of autoantibodies through molecular mimicry, dysregulated brainstem and vagal nerve signaling, activation of primed immune cells, and/or clotting/coagulation vascular issues^[8,22]^.

As of today, treatments for Long COVID are not standardized and have been ineffective^[20,23]^. One proposed strategy for treating Long COVID is to increase the body’s ability to restore immune balance by controlling inflammation with anti-inflammatory substances, such as Vitamin C, turmeric or others. Dietary supplements like vitamins and minerals, which have anti-inflammatory and antioxidant properties, have emerged as potential treatments for Long COVID^[24]^. For this reason, the aim of this double-blind study was to evaluate the supplementation of patients with EchA Marine^®^, a commercial dietary supplement based on sea urchin eggs rich in Echinochrome A in terms of its impact on health-related quality of life. To date, no reported treatments for Long COVID, using double-blind studies with nutraceuticals, have demonstrated an improvement in the quality of life of patients with this condition.

Echinochrome A is a natural compound found in the caviar, spines, and shells of certain species of sea urchins. The compound has demonstrated anti-inflammatory and antioxidant properties. The compound is the active agent of the clinically approved drugs, Histochrome^®^, used for myocardial infarction and Gistochrome used for glaucoma^[25]^. Echinochrome A is a 1,4-naphthoquinone polyhydroxylated pigment belonging to the family of polyphenols, with over 30 years of study and more than 90 scientific publications investigating its physiological, pharmacological and clinical effects^[26]^.

## Results

### Population data

A total of 46Long COVID patients were recruited to participate in this multicenter, double-blind, randomized study. Twenty-two patients received placebo and twenty-four received treatment with EchA Marine^®^. Both in the placebo and treatment groups, 60% of the patients were female, and there were no differences between groups (Chi-square= 2; p= 0.153). No significant differences were found in the Body Mass Index and age between the two groups (F_1,44_= 2.011; p= 0.163 and F_1,44_= 1.363 p= 0.249, respectively) (Figure 1).

**Figure 1.**
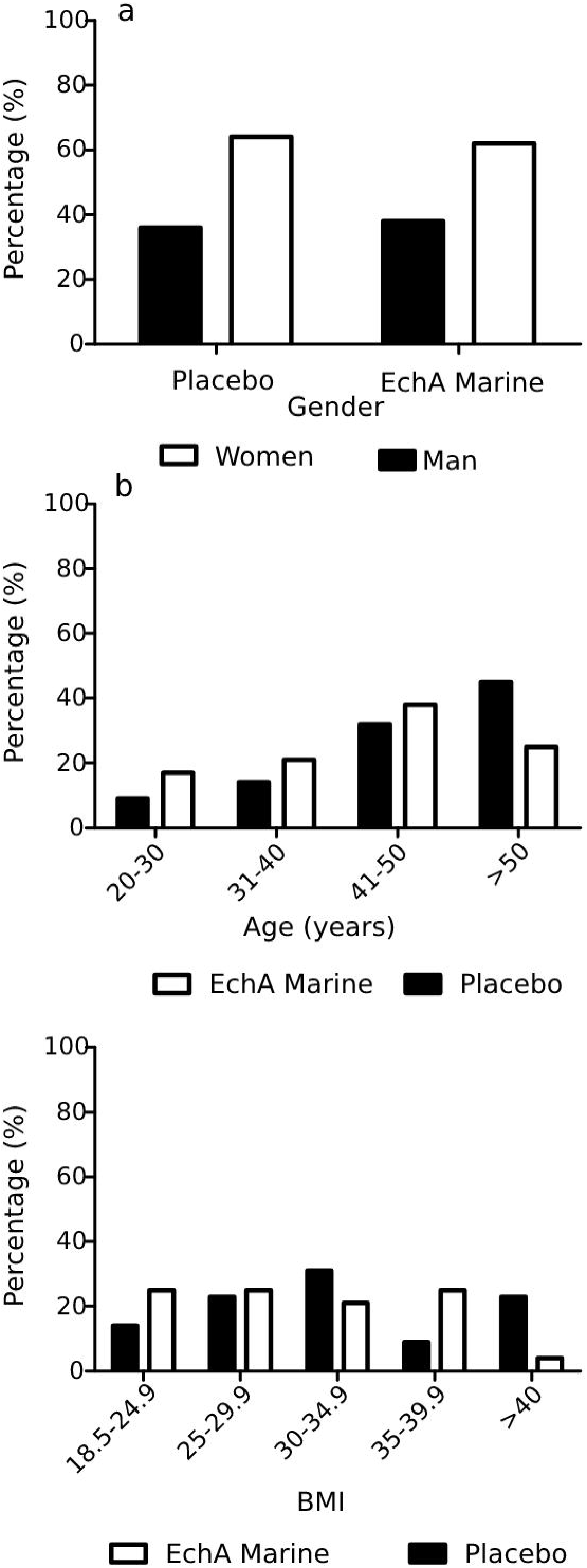
Population data. **a)** Gender. No significant differences between treatments (Chi-square= 2; p= 0.153; α=0.05). **b)** Age. No significant differences between treatments (F_1,44_= 2.011; p= 0.163; α=0.05). **c)** Body Mass Index. No significant differences between treatments (F_1,44_= 1.363 p= 0.249; α=0.05).

### Quality of life

At the beginning of the study, no significant differences were found between the groups in terms of the values in the five dimensions (Table 1). All patients reported a decrease in quality of life in some of the five dimensions, with values of 2 and 3 being the most frequent (Figure 2).

**Table 1.** ONE WAY ANOVA results summary of the EuroQol-5D. Basal condition.

|  |  | ANOVA |  |  |  |  |
| --- | --- | --- | --- | --- | --- | --- |
|  |  | Sum of |  | Root mean |  |  |
|  |  | Squares | Df | Square | F | Sig. |
| Mobility Basal | Between groups | .333 | 1 | .333 | 6.738 | .256 |
|  | Within groups | 11.080 | 44 | .252 |  |  |
|  | Total | 11.413 | 45 |  |  |  |
| Self Care Basal | Between groups | .129 | 1 | .129 | .008 | .448 |
|  | Within groups | 9.697 | 44 | .220 |  |  |
|  | Total | 9.826 | 45 |  |  |  |
| Usual Activities Basal | Between groups | .008 | 1 | .008 | 1.302 | .899 |
|  | Within groups | 21.731 | 44 | .494 |  |  |
|  | Total | 21.739 | 45 |  |  |  |
| Pain/Discomfort Basal | Between groups | .011 | 1 | .011 | 8.863 | .865 |
|  | Within groups | 15.924 | 44 | .362 |  |  |
|  | Total | 15.935 | 45 |  |  |  |
| Anxiety/Depression Basal | Between groups | .042 | 1 | .042 | .678 | .797 |
|  | Within groups | 27.697 | 44 | .629 |  |  |
|  | Total | 27.739 | 45 |  |  |  |
| VAS Basal | Between groups | .004 | 1 | .004 | 5.632 | .734 |
|  | Within groups | 1.576 | 44 | .036 |  |  |
|  | Total | 1.581 | 45 |  |  |  |
| TTO Basal | Between groups | .064 | 1 | .064 | 9.746 | .358 |
|  | Within groups | 3.269 | 44 | .074 |  |  |
|  | Total | 3.333 | 45 |  |  |  |

Significant differences $p < 0.05$ .

**Figure 2.**
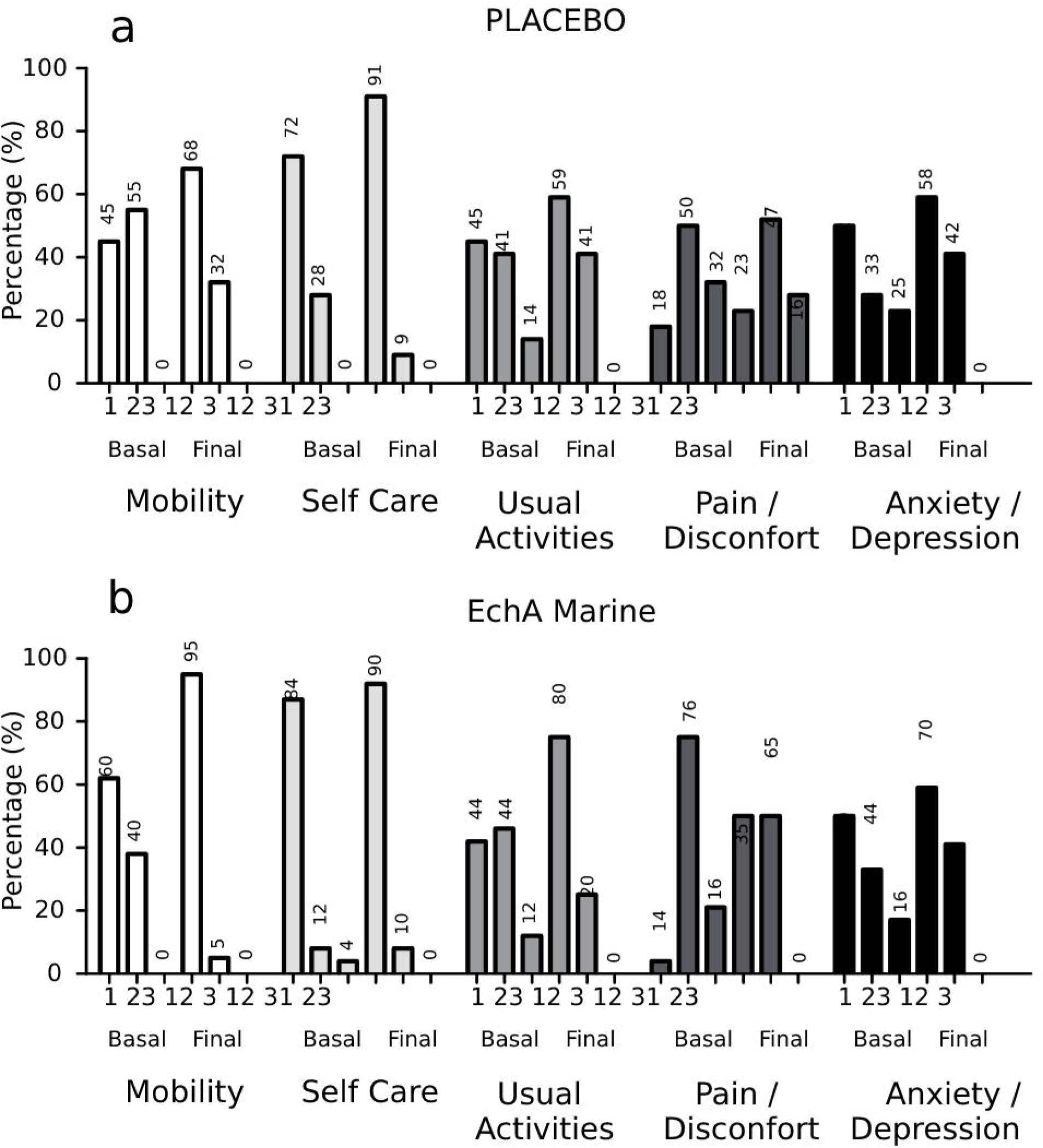
Percentage of patients in basal and final conditions for each of the five EuroQol dimensions: Mobility, Self care, Usual Activities, Pain/Discomfort and Anxiety/Depression. **a)** Placebo, **b)** EchA Marine^®^. Three-level scale for the degree of the symptom is represented by numbers in x axis with 3 meaning the least positive state and 1 the best. Mobility and Pain/Disconfort presented significant differences between treatment (F_1,44_= 6.738 p= 0.013; F_1,44_= 3.415 p= 0.005; α=0.05).

After three months of treatment, intervention with the dietary supplement EchA Marine^®^ achieved a significant decrease in the values of two of the five dimensions of the EuroQol (Figure 2, Table 2). Specifically, in mobility and pain and discomfort. The most significant effect was observed in the reduction of pain and discomfort, where the placebo did not have a significant effect. In fact, patients who consumed the dietary supplement did not present values of 3 in any dimension at the end of the treatment, while patients in the placebo group still presented values of 3, indicating a positive resolution of symptoms when consuming EchA Marine^®^. As for TTO and VAS, a significant difference was observed in both cases (Table 2), indicating an increase in general Quality of Life after treatment. These results can be observed in Figure 3.

**Table 2.** ONE WAY ANOVA results summary of the EuroQol-5D. Final condition.

| **ANOVA** |  |  |  |  |  |  |
| --- | --- | --- | --- | --- | --- | --- |
|  |  | Sum of |  | Root mean |  |  |
|  |  | Squares | Df | Square | F | Sig. |
| **Mobility Final** | Between groups | .878 | 1 | .878 | 6.738 | **.013** |
|  | Within groups | 5.731 | 44 | .130 |  |  |
|  | Total | 6.609 | 45 |  |  |  |
| Self Care Final | Between groups | .001 | 1 | .001 | .008 | .929 |
|  | Within groups | 3.652 | 44 | .083 |  |  |
|  | Total | 3.652 | 45 |  |  |  |
| Usual Activities Final | Between groups | .291 | 1 | .291 | 1.302 | .260 |
|  | Within groups | 9.818 | 44 | .223 |  |  |
|  | Total | 10.109 | 45 |  |  |  |
| **Pain/Discomfort Final** | Between groups | 3.415 | 1 | 3.415 | 8.863 | **.005** |
|  | Within groups | 16.955 | 44 | .385 |  |  |
|  | Total | 20.370 | 45 |  |  |  |
| Anxiety/Depression Final | Between groups | .158 | 1 | .158 | .678 | .415 |
|  | Within groups | 10.277 | 44 | .234 |  |  |
|  | Total | 10.435 | 45 |  |  |  |
| **VAS Final** | Between groups | .171 | 1 | .171 | 5.632 | **.022** |
|  | Within groups | 1.340 | 44 | .030 |  |  |
|  | Total | 1.511 | 45 |  |  |  |
| **TTO Final** | Between groups | .279 | 1 | .279 | 9.746 | **.003** |
|  | Within groups | 1.258 | 44 | .029 |  |  |
|  | Total | 1.536 | 45 |  |  |  |

Significant differences $p < 0.05$ .

**Figure 3.**
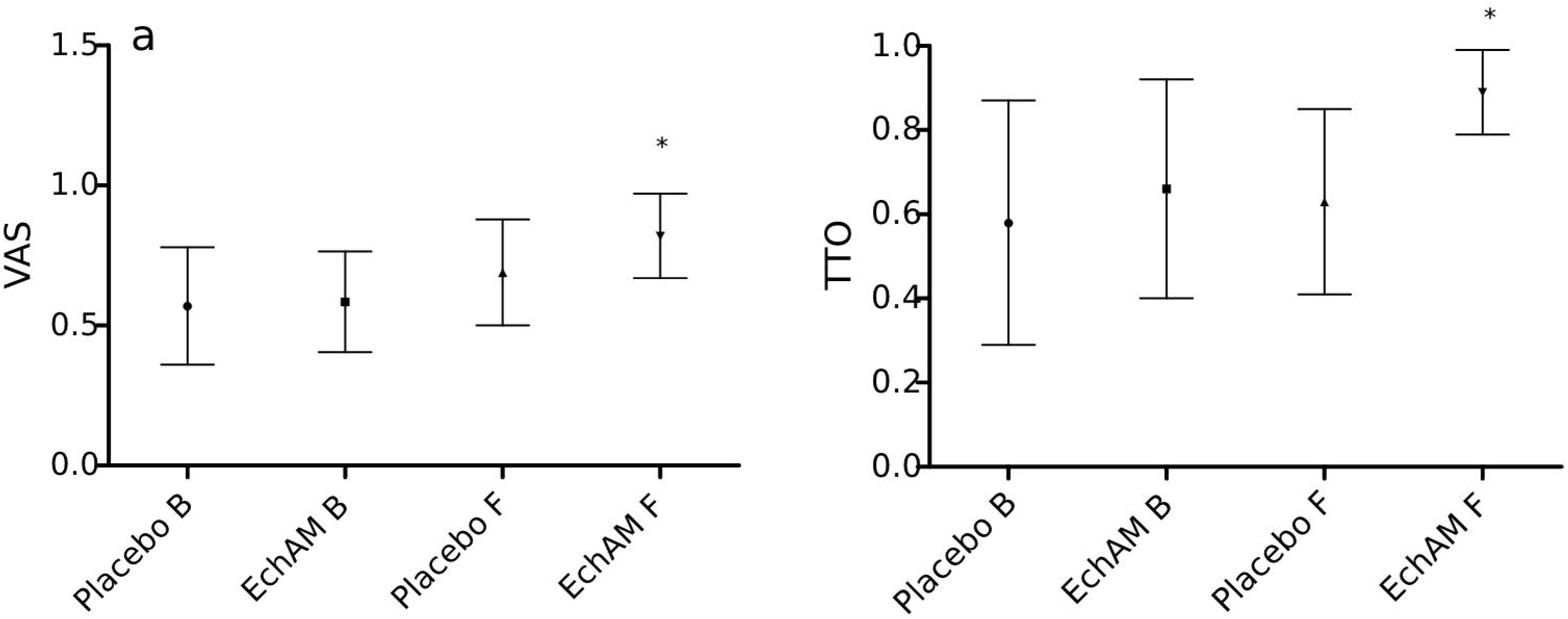
Valuing health status by VAS **(a)**, and TTO **(b)**. B: Basal, F: Final, EchAM: EchA Marine^®^. Bars represent standard deviation. Treatments presented significant differences both in Basal and Final conditions (F_1,44_= 5.632 p= 0.022; F_1,44_= 9.746 p= 0.003; α=0.05).

## Discussion

The COVID-19 pandemic has led to a significant number of cases of Long COVID syndrome, which can persist for months and substantially impact patients’ quality of life^[3]^. Treatment of Long COVID can be challenging, and existing options have had limited success^[20,27]^.

This is the first multicenter, double-blind, intervention study assessing the quality of life in Long COVID patients, and we have demonstrated that the dietary supplement EchA Marine^®^ can significantly enhance the quality of life of these patients, particularly as regards pain and discomfort; notably improving their quality of life and ability to have a better mobility. Although values were not significant in other dimensions, it can be observed that the group of patients who consumed the dietary supplement did not have values of 3 at the end of the study, while patients with placebo still had these values. This indicates that the supplement improved all dimensions studied, evidenced by the TTO and VAS values, and the prolonged consumption of the supplement may reduce values in other dimensions. Considering that, despite the small sample size, the results were statistically significant, and can be seen to be robust, offering promising implications for the treatment of Long COVID syndrome. Furthermore, this suggests that with a larger number of patients, the results could be even more solid and promising. This indicates that EchA Marine^®^ is an effective and natural treatment option for improving the quality of life of Long COVID patients. These findings are especially encouraging given the safety and natural origin of the supplement. Importantly, the prolonged consumption of the supplement may further improve values in other dimensions.

The mechanism underlying these improvements may be related to the nature of the Echinochroma A, the active ingredient in the sea urchin eggs in the dietary supplement. This compound has been suggested to alleviate the Cytokine Storm Syndrome^[28]^. The most significant effect of this molecule at cellular level is its ability to improve cellular function. This is a small molecule of 260kDa that easily enters the cell in its salt format and performs a dual function: on the one hand, the molecule inhibits free radicals, balancing oxidative stress, and on the other, it generates large amounts of hydrogen peroxide, mimicking the SOD enzyme and activating the metabolism of glutathione, thereby generating energy through mitochondrial activation and increased mass, as well as cellular oxygenation ^[26,28,29,30,31,32]^. As a result, the working hypothesis is that Echinochrome A can effectively improve cellular function and, consequently, organ function, thereby contributing to improving the quality of life of Long COVID patients. With an ever-evolving disease like COVID-19, the identification of effective treatments for Long COVID is a priority, and compounds like Echinochrome A offer a promising alternative to existing treatments.

Overall, our study demonstrates that the dietary supplement based on sea urchin eggs extract is an effective treatment option for Long COVID patients and with further research and larger sample sizes, its efficacy could be further strengthened. By offering a safe and natural solution to Long COVID, EchA Marine^®^ has the potential to make a significant impact on the fight against the pandemic and can improve the lives of those affected by Long COVID. The medical and scientific community is facing an ever-evolving disease, and the identification of effective treatments for Long COVID is a priority.

## Methods

### Participants

We carefully and prospectively selected 47 patients of both sexes, between 18 and 60 years, with the Long COVID syndrome, who came to our center (Hospital Donación Santojanni y Hospital de Agudos Ramos Mejía, City of Buenos Aires, Argentina,) between September 2021 and October 2022. (https://clinicaltrials.gov/ct2/show/NCT05531019?term=sea+urchin+eggs&draw=2&rank=1).

### Definition of Long COVID Syndrome

Long COVID Syndrome was defined as individuals who showed symptoms that had developed, during or after infection, consistent with COVID-19,which had continued for 12 weeks or more and which had not been explained by alternative diagnoses. They had had a past infection with SARS-CoV2 confirmed by PCR or antigen and who still had (at least 3) symptoms after 12 weeks: headache, dyspnoea, anosmia, ageusia, cough, chest pain, heart palpitations, abdominal symptoms, myalgia, fatigue, cognitive difficulties, depression, or anxiety. These individuals most often showed a group of symptoms, overlapping one another, and which had changed over time, affecting a number of different systems in the body^[2,3,4]^.

### Inclusion criteria

Positive diagnosis of COVID-19 in at least the last 12 weeks, diagnosis of Long COVID, adult men or non-pregnant adult women between the ages of 18 and 60, the patient provides their informed consent prior to initiating any study procedure.

### Exclusion criteria

Patients without any Long COVID symptoms or those who do not want to give their informed consent or those without a positive result for COVID-19 or with pregnancy or lactation, or those who, in the opinion of the doctor, evidence some form of an advanced organic dysfunction that would not make participation appropriate.

### Study design

The study is a prospective, randomized, double-blind, multi-hospital clinical trial with intervention.

### Demographic and clinical information

Patients also provided sociodemographic information, such as sex, age, weight, and height. They also provided relevant clinical information, such as alcohol consumption, smoking, medication, and medical history.

### Health-related quality of life (EQ-5D-3L)

Patients completed the EuroQol questionnaire in the hospital at which they were recruited during an interview with a physician belonging to the study team. These interviews were held at the beginning and at the end of the study. The questionnaire was administered in paper format. The data was subsequently uploaded to the SkyMed telemedicine platform (Ocus Cloud) to make it available for statistical analysis. The EuroQol EQ-5D-3L questionnaire was used to assess health-related quality of life and consisted of two sections: a visual analog scale (VAS) and a descriptive system covering five dimensions (mobility, self-care, usual activities, pain/discomfort, and anxiety/depression)^[33]^. Each dimension is evaluated on a three-level scale, allowing patients to be classified into 243 different health states. The scores of the dimensions have different meanings, with 3 representing the least positive state and 1 the best ^[34]^. The values obtained for Argentina in the study by Augustovski^[35]^ were applied to assign quality of life scores and used time trade-off (TTO) and contingent valuation with regression analysis (VAR) techniques. The EuroQol dimensions were measured and the TTO and VAS techniques were used to evaluate patient quality of life before and after treatment.

### Randomization

Patients were randomly assigned to each group (placebo and treatment) following a double-blind pattern. The R software was used to perform a permuted block randomization, with a sequential mixed block size of 6. Randomization was supervised by a statistics specialist who was a member of the scientific team and who was specifically designated for this purpose. This individual did not participate in any patient-related activities. This individual coordinated the delivery of the nutraceutical product to the patients. The randomization results were blinded for the remainder of the research team and for the patients. The product and placebo were optically indistinguishable.

### Treatment

Patients in the treatment group received two doses of 3ml per day of the dietary supplement EchA Marine ^®^ with a concentration of 0.025% of Echinochrome A from sea urchin roe, twice a day for 90 days. Patients consumed 1.5 mg of Echinochrome A per day. The placebo group received an equivalent solution but without Echinochrome A.

### Informed consent

Patients (or a legally authorized representative) provided their informed consent prior to initiating any study procedures.

### Study duration

The study was conducted for a total of 90 days.

### Outcome variables

The primary outcome variable will be the improvement in patients’ quality of life, analyzed on the basis of its 5 dimensions and 3 levels. The safety and efficacy of the product will also be evaluated. The study compared the efficacy of the dietary supplement Echinochrome A with a placebo in patients with Long COVID symptoms.

### Ethical considerations

The researchers participating in this project are aware of, and comply with, all ethical, legal, and regulatory safeguards established in National bioethical standards - ANMAT 5330/97 disposition - and International standards - Nuremberg Code, Helsinki Declaration version 2008 and its amendments, Universal Declaration on the Human Genome and Human Rights adopted by the UNESCO General Conference in 2005, and in accordance with the regulations of the International Declaration on Human Genetic Data UNESCO 2003, the Universal Declaration on Bioethics and Human Rights, and the Guide for Human Health Research (GISH), resolution 1480/2011 MSN. This study has been approved by the Ethics Committees of the Santojanni, Ramos Mejia, and Muñiz hospitals. All detailed information regarding this medical trial can be found on ClinicalTrials.gov Identifier: NCT05531019.

### Data analysis

Data was downloaded from the OcusMed platform into Excel spreadsheets and then analyzed using the SPSS statistical software version 25. A Chi-Square analysis was performed for gender differences between treatments. One-way analysis of variance was used for age and BMI differences between treatments. Quality of life scores were calculated for each dimension of EuroQol and for the VAS scale. One-way analysis of variance was used to compare quality of life scores between patients in the treatment group and the placebo group. A p-value <0.05 was considered statistically significant.

### Confidentiality of Information

In Argentina, there is a law protecting personal information (Law No. 25,326 on Personal Data Protection) which establishes the integral safeguarding of data recorded in files or records, whether they be public or private in nature, to guarantee the right to privacy of individuals. When referring to “Consumers personal data’’, the Law refers to the personal data controlled by authorized institutions and which OcusCloud processes in the course of providing the SaaS (Software as a Service) SkyMeD. The terms “controller”,” “processor,” “processing,” “processed,” “processing,” and “personal data” are defined according to the European Union Directive 95/46/EC, except as defined by corresponding data protection legislation in Argentinean Law. OcusCloud guaranteed, on the basis of a confidentiality agreement, the implementation of the necessary technical and organizational measures to protect the data provided in all means/systems under its control and supervision. There is also Law No. 26,529 on Patient Rights, in force since February 2010, which regulates civil relationships between patients and doctors and health institutions and legislates on the information that doctors must provide and patients must receive in terms of clinical documentation. The patient’s clinical information and documentation are adequately safeguarded for their privacy, as well as regards the confidentiality of sensitive data. This includes statistical confidentiality in relation to the development of databases in the health system, whereby individual statements and/or information cannot be communicated to third parties, nor used, disseminated, or published in a manner allowing the identification of the person or entities. Sensitive data related to the patient is protected by confidentiality and privacy, as well as data arising from research. Only investigators directly involved in the trial and members of the Ethics Committees of the participating hospitals will have access to this information.

### Interview description

EuroQol surveys were undertaken face-to-face at the start of the study and, again, after three months. Day 1 is the day of the first intake of EchA Marine ^®^. Once the patient had been included in the study, the objective and design of the study were carefully explained and a brochure with information regarding the clinical trial and the nutraceutical was provided to the patient. It was mandatory that the patient formally confirm their acceptance of the express informed consent by signing the informed consent.

## Data Availability

The datasets generated and/or analysed during the current study are available in the CONICET Institutional repository.

https://ri.conicet.gov.ar/handle/11336/199122

## Acknowledgements

This work was founded by the grant “PICTO Sequelas COVID-19 00009” from The National Agency for the Promotion of Research, Technological Development and Innovation. EchA Marine^®^ supplements for the complete clinical trial were donated by Erisea S.A.

## Author contributions

BV: Performed recruitment of patients and EuroQol interviews at Santojanni Hospital and writing of the manuscript

RT: Performed the analysis of the data and writing of the manuscript TJ: Performed recruitment of patients

MP: Performed recruitment of patients

CAA: Performed data analysis

BE: Performed data analysis

dRN: Performed recruitment of patients

IVJ: Performed recruitment of patients

JN: Performed recruitment of patients

VC: Design, graphics and format of the paper

SM: Medical responsible at Ramos Mejia Hospital. Revision of manuscript

dLG: Protocol Design and data analysis. Medical responsible at Muñiz Hospital. Revision of manuscript

SF: Medical responsible at Santojanni Hospital. Revision of manuscript

## Additional Information

### Competing interests

TR is co-founder of Erisea S.A. that produced the Dietary Supplement for the Clinical Trial. VC is employee of EriSea S.A. The rest of the authors declare no potential conflict of interest.

## Data availability statement

The datasets generated and/or analysed during the current study are available in the CONICET Institutional repository. [https://ri.conicet.gov.ar/handle/11336/199122]

## Notes

### Clinical Trial

NCT05531019

### Author Declarations

This study has been approved by the Ethics Committees of the Hospital Donacion Francisco Santojanni (CR 5832), Hospital General de Agudos Jose Maria Ramos Mejia (CR5196), and Hospital de Infecciosas Francisco Javier Muniz (CR 5240).

